# Digital therapy to improve stuttering symptoms in patients with a deficit of spectral power in the EEG beta frequency band

**DOI:** 10.1101/2022.04.21.22272785

**Authors:** Dmytro Chernetchenko, Pramax Prasolov, Sam Aganov, Andrey Voropay, Dmytro Lituiev, Eugene Nayshtetik

**Affiliations:** ZOV.ai, Research and Development Lab, Kyiv, Ukraine and San Francisco, CA, USA; Dnipro National University, Department of Physics, Electronics and Computer Systems, Dnipro, Ukraine; UC Berkeley, ZOV.ai, San Francisco, CA, USA; California Institute for Human Science, Encinitas, CA, USA; University of the Cumberland, Williamsburg, KY, USA; UCSF, San Francisco, CA, USA

**Keywords:** speech disorders, binaural beats, auditory stimulation, EEG patterns, stuttering

## Abstract

**Background:** Stuttering is a speech disorder that affects more than 70 million people worldwide, limiting their ability to communicate and socialize. In recent decades, several studies have demonstrated a link between stuttering and a deficit of *β* electroencephalographic (EEG) power.

**Aim and Methods:** This study investigates the efficacy of a novel auditory neuro modulating technology that leverages euphonic music tracks with broad-spectrum binaural beats to induce selective EEG spectral power changes. Adults with stuttering (AWS, *n*=6) and participants from the control group (*n*=6) were exposed to euphonic binaural stimuli for 5 minutes. The EEG and electrocardiographic (ECG) bio-signals were recorded prior, during, and after exposure.

**Results:** During standard reading tasks without stimulation, *β*-power in the left hemisphere in the adults without stuttering and with stuttering differed. The left-right hemisphere asymmetry in *β*-wave power was observed in the control group but not in AWS. After the stimulation, the power of *β*-band in AWS participants in the left hemisphere increased 1.54-fold, while changes in the right hemisphere activity were not significant. Average *β*-band power within left frontotemporal area and temporoparietal junction after stimulation in AWS participants shows an increase of *β*-band in left frontotemporal junction by 1.65-fold and in left temporoparietal by 1.72-fold. The changes in the quality of speech were assessed based on the speech rate and the rate of speech disfluencies evaluated by speech therapists. The rate of disfluencies dropped significantly after the stimulation (median 74.70% of the baseline rate), but the effect was not significantly different from the baseline 10 min later (median 65.51% of the baseline rate). Similarly, the speech rate significantly increased immediately after the stimulation (median 133.15%) but was not significantly different 10 min later (median 126.63% of the baseline rate). In this study, we found significant correlations of *β*-activation level in left temporoparietal projection (Spearman *ρ*=-0.54,) and left frontotemporal area (Spearman *ρ*=-0.58) with disfluency rate of speech.

**Conclusions:** We show for the first time that auditory binaural beats stimulation can substantially improve speech fluency in AWS and its effect is related to boost of EEG *β*-band power in speech-production centers. The changes in *β* power are detected immediately after the exposure and persist 10 min later. Additionally, these effects are accompanied by a reduction in stress level as monitored by ECG markers. This suggests that auditory binaural beats stimulation temporarily improves speech quality in AWS by increasing *β*-band power of EEG in speech centers of the brain.

## BACKGROUND

Stuttering is a speech disorder that affects 2.8–3.4% of children at some developmental stage. Even though 80% of children recover, both with natural development and therapy, the other 20% continue to stutter in their adulthood, which accounts for 1% of the adult population^1,2,3^. Causes of stuttering are still poorly understood but are usually attributed to speech motor control disruption due to abnormal electrical activity of the brain, anatomical abnormalities, genetic and molecular factors, stress or psychological trauma, or developmental delay. Neurophysiology studies have offered a promise of actionable discoveries in stuttering causes. A number of studies showed that the activity in the EEG *β* spectrum range (13-30 Hz), which is commonly heightened in awake state and focused activities^4^, is reduced in individuals with stuttering (reviewed in ^5^). EEG and magnetoencephalographic (MEG) studies have shown that in adults who stutter (AWS), as compared to the control group, *α* power is higher ^5,6^ while *β* power is lower during speech production^6^. This may be due to the role of *β* band in coordinating speech planning and execution within the speech-production brain network, as it is known that *β* oscillations become more coherent between the bilateral primary motor, premotor, and auditory cortices before speech production^7^. Additionally, EEG and MEG studies suggest that coordination between auditory clues and motor excitation, which is carried out through *β* oscillations^8^, is affected in AWS ^9,10,11^.

Several methods have been developed to entrain brain oscillations. Synchronization of brain activity at the frequency of sensory stimulation and its harmonics, in and beyond the corresponding sensory brain areas, has been achieved with visual flicker^12^, somatosensory tactile stimulation^13^, and auditory stimulation. For auditory brain entrainment, popular approaches include isochronous sounds^14,15^, monaural beats ^16,17,18^, and binaural beats ^18,19,20^. Stuttering reduction using noise and auditory voice feedback treatment has been explored for several decades^21,22^. Still, a recent advancement is a targeted enhancement of the *β* band through sound waves. Feasibility of entraining *β* band by isochronous sound exposure has been shown in healthy adults^15^ and in AWS^23^.

The earlier studies found that the brain activity patterns normally seen during speech production in non-stutterers were either absent, bilateral in nature, or lateralized to the left hemisphere. Often, in previous works underlines a high impact^24^ of the auditory pathway which provides the normal and synchronic activation in speech areas of the brain during fluent speech production.

Given the earlier results suggesting possible connection between stuttering and the deficit in spectral power of *β* EEG band^25,26,15^ and the lack of activation^27^ in speech production centers of the brain, we hypothesized that increase of *β-*power with binaural beat stimulation might temporarily alleviate disfluency episodes and improve speech quality in AWS.

To test this hypothesis, we exposed people with stuttering (AWS) and fluent individuals (controls) to proprietary binaural beat soundtracks. Simultaneously, we recorded EEG, ECG, and speech production. To reveal the influence of the auditory stimulation on electrophysiological brain activity, we analysed the whole spectrum of EEG and performed separate analysis for the electrodes group located over the left frontotemporal (LFT) area and temporoparietal junction (LTP).

## MATERIAL AND METHODS

### Experimental design

Adult volunteers with stuttering (AWS, 6 males; aged 18–38 years; mean age: 28.3 ± 7.1 years) and fluently speaking volunteers (controls, 6 males; 26–52 years old; mean age: 32.0 ± 10.1 years) signed informed consent and participated in the study at the laboratory of Faculty of Physics, Electronics and Computer Systems of Dnipro National University (study approved by the Ethical Commission decision, protocol #15 from 2021-05-17). All participants were right-handed. All control participants reported no history of speech, language or hearing difficulties. Experimental protocol consisted of six stages, 5 min each (Fig. 1A): (1) baseline exposure under relaxation and with eyes closed, (2) reading activity pre-stimulation, (3) auditory stimulation, (4) reading task activity right after the stimulation, (5) reading task activity after 10 minutes of relaxation, and (*6*) resting state measurement with eyes closed. During each stage, EEG and ECG signals were continuously recorded. Reading aloud was assayed in participants’ native language (Russian) for 5 min. Voice was recorded with a built-in microphone of a MacBook Pro 13” (2015) laptop.

**Figure 1.**
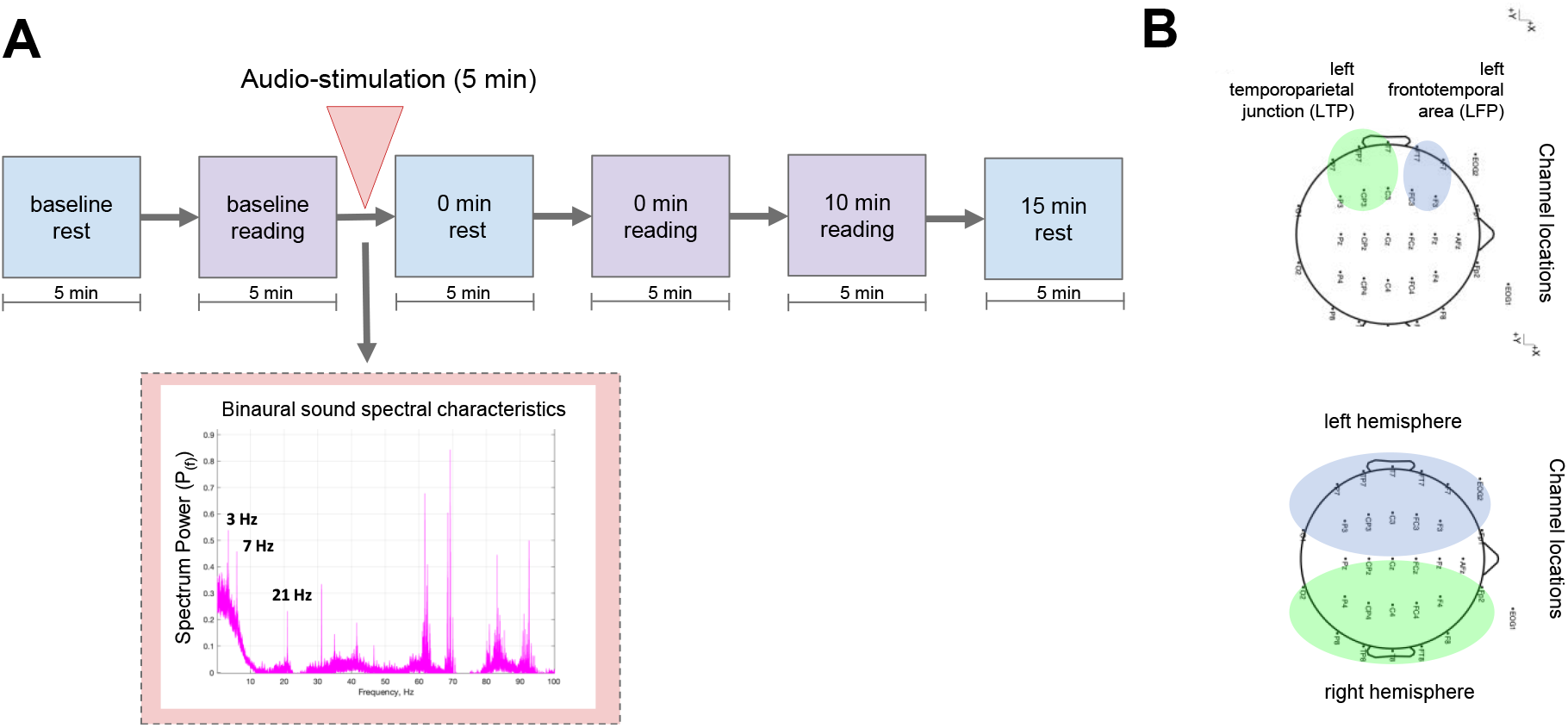
The design of the study. **A**. Scheme representing the design of the experiments. **B**. Location of EEG electrodes (32 in total) used during all experiments with highlighted groups of electrodes over the left temporoparietal junction (green on upper topography) and left frontolateral area (blue on upper topography) and groups of electrodes used for the whole-hemisphere analysis (bottom topography, blue for the left and green for the right hemisphere). Abbreviations: EEG - electroencephalographic

**Figure 2.**
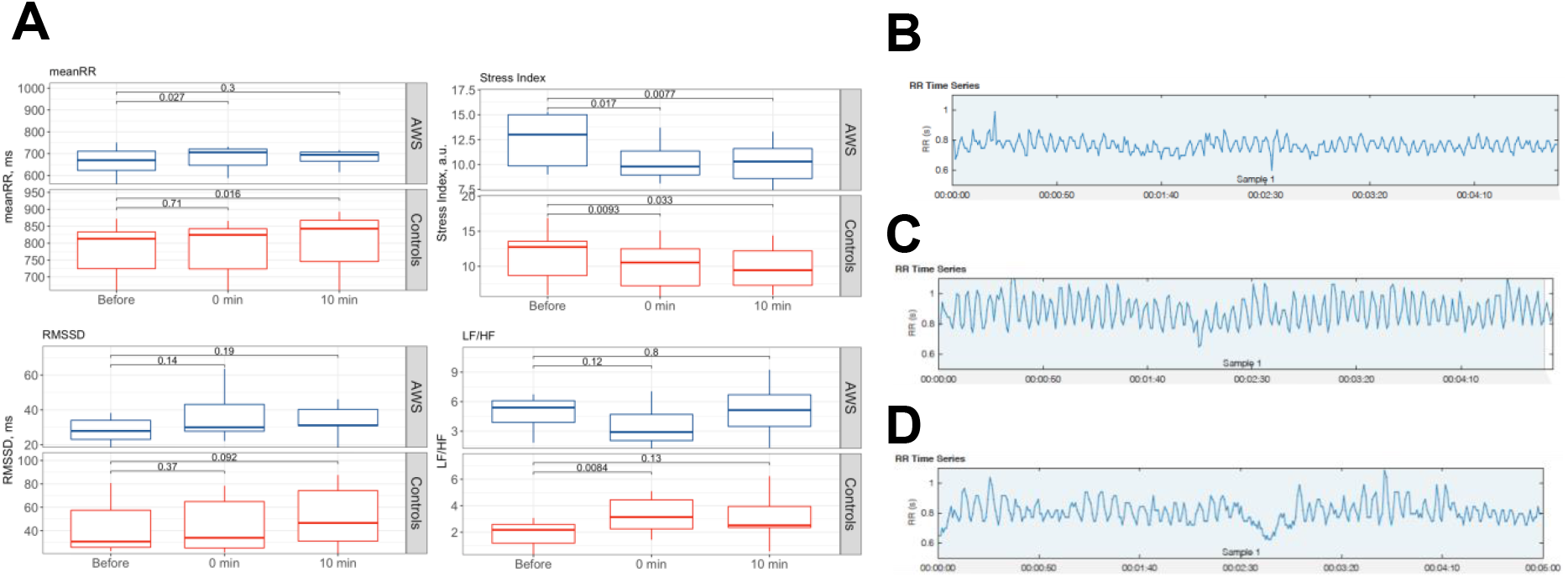
ECG-based markers of stress during reading tests. **A**. Four markers of HRV (mean RR, Stress index, RMSSD, LF/HF) are shown for AWS and control group before, immediately after (0 min), and 10 min after the binaural beats stimulation. RR intervals deviation with time demonstrate HRV characteristics before stimulation (**B**), right after stimulation (**C**), and 10 min after stimulation (**D**) Abbreviations: ECG – electrocardiography, HRV – heart rate variability, AWS – adults with stuttering; RMSSD – root mean square of successive differences, LF/HF – low-to-high frequency ratio

### Auditory Stimulation

An auditory euphonic binaural beat (BB) stimulus with three spectral components (*δ, α*, and *β*) was prepared using a custom generator and post-processing algorithms [United Kingdom Patent Application No. 2116927.1]. Music with vocals was used as a carrier to target brain cortical centers responsible for voice recognition specifically. To produce a euphonic binaural stimulus^28^, we applied a low-pass filter with cut-off of 170 Hz, which was derived empirically for chosen music background. The resulting track was normalized at the final stage, and an output stereo audio raw data was generated. The stimulus used in this study contained binaural components at 3 Hz, 7 Hz and 21 Hz with a high-pass cut-off of the binaural effect at 170 Hz (see Supplementary Fig. 1A and Supplementary Fig. 1B). Spectral characteristic of the resulting output auditory stimulus is shown in Supplementary Fig. 1C. The output intensity of the stimulus was normalized to 55 dB level to match the optimal level of auditory stimulation^29^. Auditory stimulus was delivered using high-quality wireless *Sennheiser momentum true wireless 2* headphones with an output frequency response band in the range of 5 to 33,000 Hz.

### EEG registration and signal processing

A medical-grade digital brain electric activity monitor *(CONTEC KT88-3200*, S/N 591012399249) was used to collect EEG raw data with conventional wet cap electrodes. Raw data collected with proprietary companion software tool CONTEC EEG32. Output raw data was saved into the European EDF+ format. We used 10/20 EEG electrode placement according to the international standard system^30^, with reference electrodes A1 and A2 (the electrodes configuration shown on Fig. 4A).

**Figure 3.**
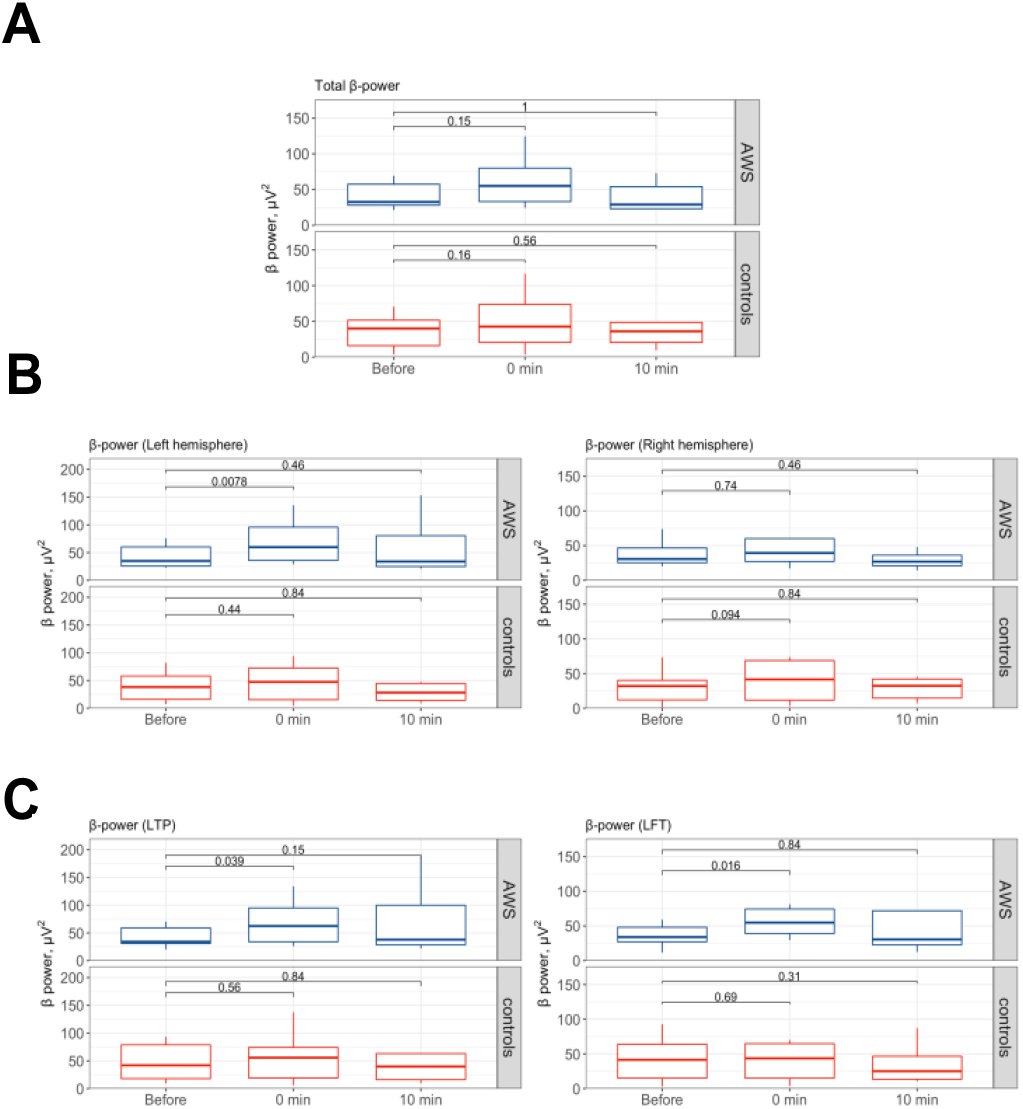
EEG *β*-power change at the different stages of experiment. **A**. EEG *β*-waves in reading experiments in AWS (top) and control group (bottom) averaged across all 32 electrodes. **B**. Power in *β*-band for left and right hemispheres (left and right, respectively) during the reading activities in AWS and controls. **C**. Power in *β*-band averaged across electrode groups located over the LTP junction (left) and LFT area during the reading activities in AWS and controls. Abbreviations: EEG – electroencephalography, AWS – adults with stuttering; LTP – left temporoparietal junction; LFT – left frontotemporal area

**Figure 4.**
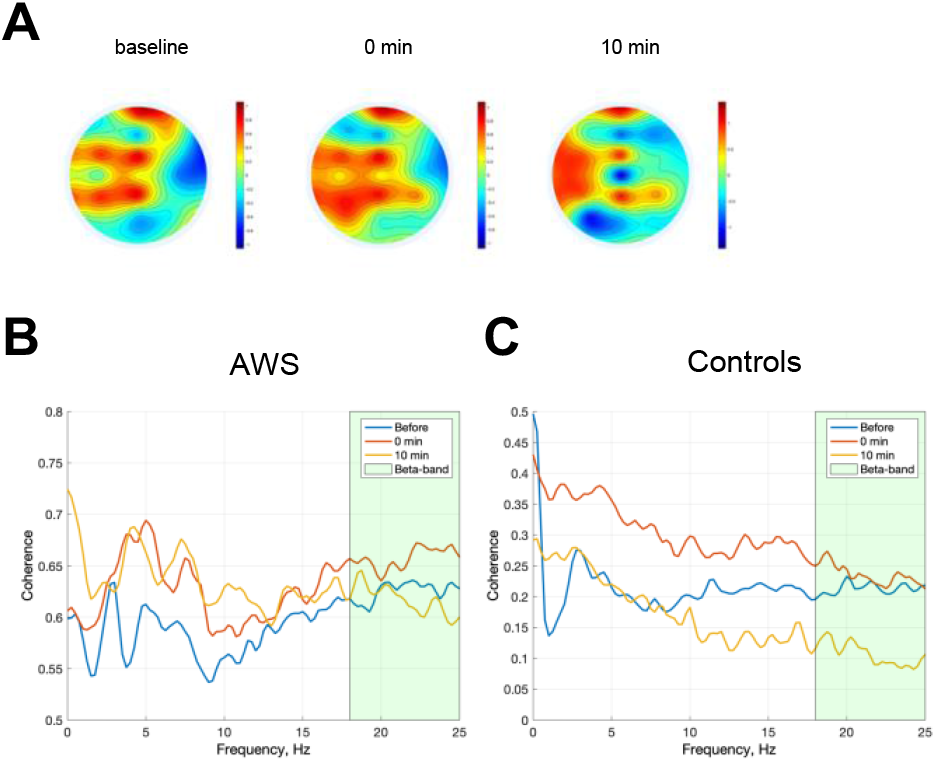
Topography and frequency specificity of binaural beats stimulation-induced changes. **A**. Topographical distribution of *β-* power before, after and 10 minutes after the stimulation. **B**. Coherence between LTP junction and LFT area during speech in AWS, before, immediately after and 10 min after stimulation. **C**. Coherence between LTP junction and LFT area during speech in controls, before, immediately after and 10 min after stimulation Abbreviations: AWS – adults with stuttering; LTP – left temporoparietal junction; LFT – left frontotemporal area Abbreviations: AWS– adults with stuttering; LTP – left temporoparietal junction; LFT – left frontotemporal area

Sampling rate for each channel was 200 Hz with 0.1 µV resolution and a 0.016–1000 Hz input signal frequency range. The input data were digitally filtered with a *f*=0.1–30 Hz band pass filter to exclude electromyographic (EMG) artefacts. Brain electrical activity was postprocessed using Fast Fourier Transform (FFT) followed by power spectrum density (PSD). EMI noise was reduced by applying a notch filter with a cut-off frequency of 50 Hz. Episodes of electrodes lead-off were excluded from the record by an automatic built-in lead-off detection function of EEG monitor. Additionally, EOG artifacts are extracted using Independent Component Analysis (ICA) Infomax decomposition algorithm. Absolute power was numerically integrated over each frequency of the following bands: *δ* (0.5 – 4 *Hz*), *θ* (4 – 7 *Hz*), *α* (7 – 13 *Hz*), and *β* (13 – 30 Hz) over every electrode’s position (32-electrode EEG measurement). Consequently, the average of 32 channels was used for statistical analysis of spectral power. Spatial distribution of PSD (calculated using a standard formula^29^) was visualized as heatmaps for each EEG spectral band. To obtain the power of electrical brain activity in areas associated with speech production processes we calculate the mean average power at left temporoparietal junction (anatomically associated with Wernicke’s area) within electrodes C3, CP3, P3, P7, TP7, T7 and left frontotemporal area within electrodes F3, FC3, F7, FT7 (anatomically associated with Broca’s area). Topographies, compressed spectrum graph, and trend graph were produced using open-source EEGLAB v.2021.1 software.

### ECG registration and signal processing

Electrocardiographic (ECG) recordings were conducted using a *CONTEC 8000GW* device (HW S/N: 39124881923) with four wet electrodes that were placed on the limbs to obtain a conventional six lead ECG system (*I, II, III, avL, avR* and *avF*). To remove artefacts, ECG lead-off events were detected and removed using built-in AC-current lead-off detection, the signal was high-pass filtered at *f*=30 Hz to prevent aliasing and exclude electromyographic (EMG) artefacts, baseline drift was removed with a low-pass filter at *f*=0.67 Hz. Electromagnetic interference (EMI) influence was reduced by applying a notch filter with a cut-off frequency of 50 Hz. Raw ECG data was exported to the HL7 aECG standard XML format.

The state of the autonomic nervous system (ANS) and the stress level were assessed by heart rate variability (HRV) using a set of standard ECG metrics set forth by the European Society of Cardiology and North American Society HRV Standards^31^: meanRR, meanHR, SDNN (standard deviation of NN intervals), RMSSD (root mean square of successive differences), LF/HF (LF/HF – low-to-high frequency ratio), and Baevsky’s Stress Index. Baevsky’s stress index (SIdx) was computed according to the formula^32^:

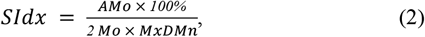

where *AMo* is the so-called mode amplitude presented in percentage points, *Mo* is the mode (the most frequent RR interval) and *MxDMn* is the variation scope reflecting the degree of RR interval variability. The mode *Mo* is simply taken as the median of the RR intervals. The *AMo* is obtained as the height of the normalised RR interval histogram (bin width 50 ms) and *MxDMn* as the difference between longest and shortest RR interval values. In order to make *SIdx* less sensitive to slow changes in mean heart rate (which would increase the *MxDMn* and lower *AMo*), the very low frequency trend is removed from the RR interval time series by using the smoothness priors method^33^.

### Speech quality assessment

Speech recordings were evaluated by speech therapists for a range of disfluencies, such as repetitions of syllables, whole words, sound prolongations, and blocks. Each audio recording was scored by a speech therapist blindly (a participant ID was provided, but not experimental stage). Disfluencies were measured in terms of the number of repetitions (for sounds, syllables, and words separately), prolongations, and blocks. Total number of disfluencies per experiment was normalized by the total number of words read. Since the texts differed from experiment to experiment, it is necessary to calculate the relative number of disfluencies for each measurement. The speech rate (words read per minute) was normalized per unit of time.

### Statistical analysis

To examine the effect of the cohort and treatment on physiological read-outs, while accounting for repeated experiments within participants, we applied linear mixed effects models (LMM)^34^. The effects due to participants’ individual physiology were treated as a varying intercept random effect, effects due to experiment were treated as random effects nested within the participants’ effects, and effects due to the cohort and or treatment as fixed effects. Analysis was performed with lme4 and lmerTest libraries in R software and significance of the fixed effects was reported. Speech characteristics were compared using Wilcoxon t-test on a single (first) experiment per participant.

## RESULTS

### Auditory stimulation results in reduction of stress level as monitored by ECG

In accordance with results of previous study^36^ where influence of binaural stimulus on the ANS were shown, we evaluate the effect of the auditory stimulation on the general stress level, heart rate and HRV parameters. Significant difference was observed for heart rate immediately after the stimulation compared to the baseline (*p*=0.024 in Wilcoxon test). After 10 min, heart rate decreased significantly only in the control group (*p*=0.01 in Wilcoxon test). However, the Baevsky’s stress index decreased in both AWS and control group relative to the baseline immediately after the exposure (by 1.16 ± 0.12 *p*=0.01 in Wilcoxon test and 1.26 ± 0.19-fold, respectively, *p*=0.03 in Wilcoxon test) and was significantly down 10 min later (*p*=0.01 and *p*=0.03 in Wilcoxon test, see Fig. 2A). No significant difference was found in HRV RMSSD after the stimulation. Examples of ANS modulation during the experiment shown by RR intervals series for one AWS participant before (see Fig. 2B), immediately after (see Fig. 2C), and 10 min after stimulation (see Fig. 2D).

### AWS display distinct spatial signatures of brain activation during speech production

To establish the validity of the method, as a first step we compared brain electric activity in AWS and control participants. No significant difference of *β* power averaged across all electrodes, neither its asymmetry was found between the control and the AWS cohort in the baseline rest state (*p*=0.43 in Wilcoxon test, see Supplementary Fig. 2A and Supplementary Fig. 2B). The total power in *β* band in the control group was significantly higher in the left hemisphere (*p*=0.03 in Wilcoxon test) during the baseline state measurements. But the electrical activity in AWS during the rest state in the left hemisphere and right hemisphere was the same (*p*=0.38 in Wilcoxon test).

Similarly, during the reading task in baseline state, average power of *β*-band did not significantly differ between AWS and the control group (*p*=1.0 in Wilcoxon test). The left-right hemisphere asymmetry in *β*-band power was observed in the control group (*p*=0.03 in Wilcoxon test) but not in AWS (*p*=0.74 in Wilcoxon test). Detailed results of hemispheres electrical activity distribution during experiment are shown on Supplementary Fig. 2A and Supplementary Fig. 2B and in Supplementary Table 1.

The *β* power in LTP junction and LFT area during the reading task did not significantly increase neither in AWS (*p*=0.64 in Wilcoxon test, *p*=0.54 in Wilcoxon test) nor in the control group (*p*=0.06 in Wilcoxon test in controls, *p*=0.15 in Wilcoxon test in controls) relative to the baseline rest state. However, the relative average power of *β*-band in LTP junction and LFT area correlated across participants in the control group (Spearman *ρ*=0.89, *p*=0.03, Supplementary Fig. 2B) as well as in AWS (Spearman *ρ*=0.9, *p*=0.05, see Supplementary Fig. 6A, Supplementary Fig. 7A).

### Auditory stimulation enhances *β* spectral power and leads to spatial redistribution of brain activity

To assess the effectiveness of the auditory binaural beat stimulation to enhance target frequencies of brain electric activity, we recorded and analysed EEG activity before, during (see Supplementary Fig. 4A), and after stimulation, in rest and during reading activity as shown in Fig. 1 and analyzed the overall power of the *β* band, as well as each hemisphere, and within speech centers projections.

The overall absolute *β* power in the control group insignificantly increased by 1.4-fold after stimulation (*p*=0.16 in Wilcoxon test) and stayed at 1.2 fold 10 min after compared to the baseline reading (*p*=0.56 in Wilcoxon test). The overall *β*-power in AWS insignificantly increased by 1.32-fold after stimulation (*p*=0.15 in Wilcoxon test) and remained at 1.14 fold 10 min after (*p*=1.0 in Wilcoxon test, see Fig. 3A) compared to the baseline reading.

Detailed analysis of spatial distribution showed that after the stimulation, the power of *β* band in AWS participants in the left hemisphere increased 1.63 fold (with *p*=0.01 in Wilcoxon test), while changes in the right hemisphere activity were not significant (*p*=0.74 in Wilcoxon test, see Fig. 3B). In the control group, no significant changes in *β* power were observed in either brain hemisphere after the stimulation (*p*=0.44 in Wilcoxon test for left, and *p*=0.09 in Wilcoxon test for right, see Fig. 3B). Spatial changes were visualized as difference topography of grand-averaged normalised *β* brain activity during the text reading after stimulation (see Fig. 4A).

Next, we analysed average *β* band power within LFT and LTP junction. After the stimulation, the average power of *β*-band within LTP projection in AWS participants increased by 1.74 fold after stimulation (*p*=0.03 in Wilcoxon test) and stayed at 1.65 fold 10 min after (*p*=0.15 in Wilcoxon test compared to the baseline). After stimulation, the average power of *β*-band within the LFT area in AWS participants increased by 1.72 fold after stimulation (*p*=0.01 in Wilcoxon test) and stayed at 1.61 fold 10 min after (*p*=0.84 in Wilcoxon test compared to the baseline). Coherence analysis between the electrodes electrical activity associated with LFT area and LTP junction in AWS cohort shown that average coherence within *β* band before stimulation was equal to 0.628 ± 0.009 (I=64.44, I_beta_=7.54), after stimulation 0.66 ± 0.01 (I=60.73, I_beta_=7.91) and 10 min after 0.606 ± 0.01 (I=62.91, I_beta_=7.27) (see Fig. 4C). At the same time, in the control group the coherence within *β* band before stimulation was 0.21 ± 0.01 (I=21.29, I_beta_=2.61), after stimulation 0.23 ± 0.01 (I=32.23, I_beta_=2.75), and after the next 10 min 0.1 ± 0.01 (I=22.64, I_beta_=1.24) (see Fig. 4D). Additionally, we observed cross-sectional correlation between activity the two areas in both AWS (see Supplementary Fig. 6A) and control (see Supplementary Fig. 6B) groups immediately after the stimulation (Spearman *ρ*=1.0, *p*=5e-5 at AWS and *ρ*=0.94, *p*=0.01 in controls respectively) and 10 minutes later (Spearman *ρ*=0.86, *p*=0.01 in AWS and *ρ*=0.97, *p*=0.001 in controls, respectively). Cross-sectional correlations for each patient shown on Supplementary Fig. 7.

### Auditory stimulation leads to improvement of speech quality conditional β-activation in speech centers in AWS

To investigate the effect of the auditory stimulation on the speech quality, we recorded speech samples before and after the auditory stimulation (as indicated in Fig. 1). The speech rate (words per minute) and the rate of disfluencies (the ratio of disfluencies to the total number of words uttered) were counted in the AWS group by a speech pathologist. The rate of disfluencies dropped after the stimulation (median 74.70% of the baseline rate, 90% CI: 37.11%–98.16%, *p*=0.04 in paired Wilcoxon test, see Fig. 5A) but there was not significant difference from the baseline 10 min later (median 65.51% of the baseline rate, 90% CI: 39.15%–93.70%, *p*=0.15, see Fig. 5B). Similarly, speech rate increased immediately after the stimulation (median 133.15% of the baseline rate, *p*=0.04), but was not significantly different 10 min later (median 126.63% of the baseline rate, *p*=0.08). Immediately after stimulation, 6 out of 6 participants noted subjective ease of reading the text aloud. Speech recordings show a smoother, clearer, and faster text pronunciation for AWS after stimulation (see the link on records in Supplementary Materials).

**Figure 5.**
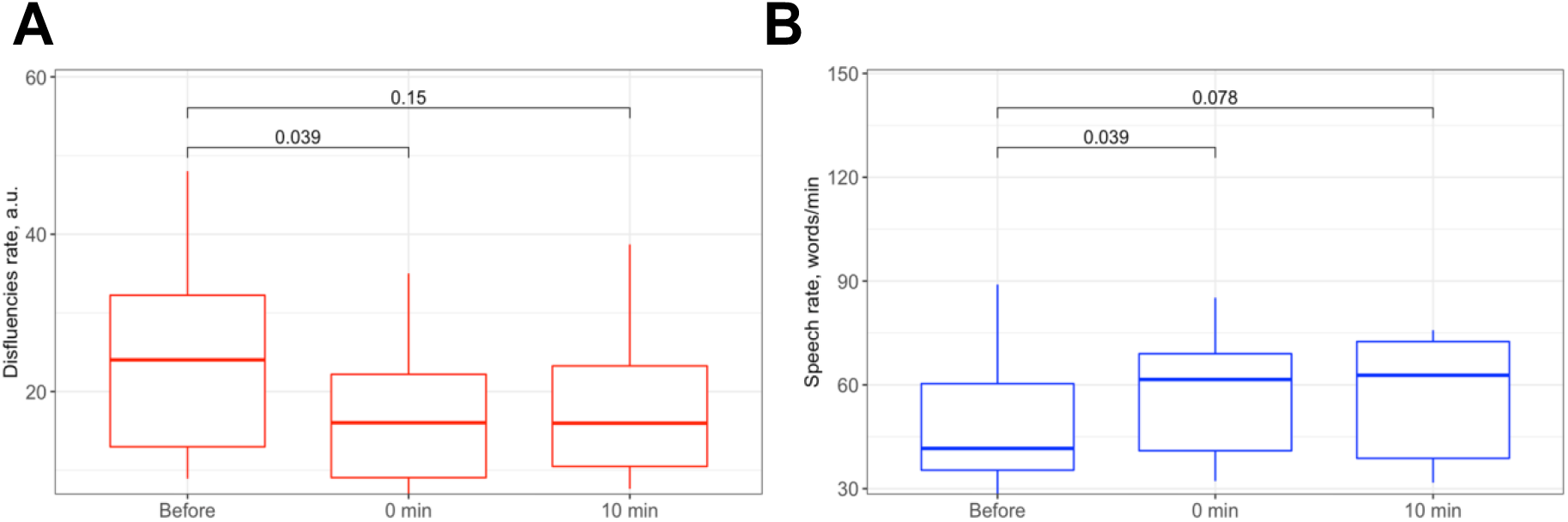
Speech disfluency and speech rate during experiments. **A**. Speech disfluency rate before, after and 10 min after binaural beats stimulation. **B**. Speech rate before, after and 10 min after binaural beats stimulation.

### Conditional improvement of speech quality related to the specific brain activity patterns

As the speech improvement is expected to be mediated by activation of LFT area and LTP junction, we analysed correlation of changes in the rate of disfluencies to activation in the respective areas. In order to elucidate the contribution of the auditory stimulation induced changes in electrical activity to changes in speech fluency, we compared changes in *β* band power in LFT and LTP junction before and after stimulation to the disfluency rate. Improvement of fluency after stimulation and 10 minutes after compared to the reference reading stage correlated with an increase in the power of *β* band in both LTP junctions (Spearman *ρ*=-0.54, *p*=0.03, see Fig. 6A) and LFT area (Spearman *ρ*=-0.58, *p*=0.02, see Fig. 6B).

**Figure 6.**
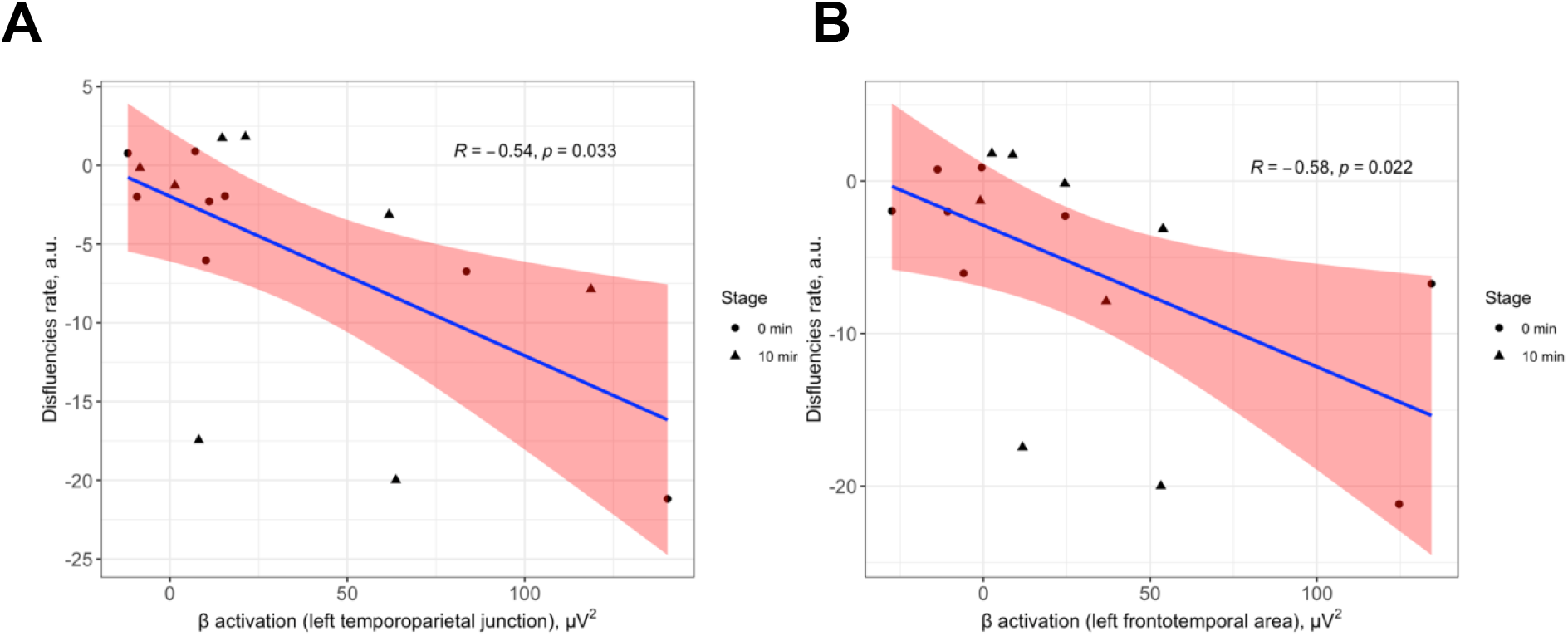
Correlation between speech disfluencies and β-power over the speech-relevant centers after binaural beats stimulation (0 min and 10 minutes after). **A**. Correlation between disfluency rate per AWS participants and β-band power average across electrodes over the LTP junction. **B**. Correlation between disfluency rate per AWS participants and β-band power average across electrodes over the LFT area.

When comparing speech fluency between the AWS participants who responded with an immediate increase of *β*-power within the speech centers upon auditory stimulation (*n*=4, average increase by 2.3 fold) to the rest AWS participants (*n*=2), the former displayed an improvement in speech fluency both immediately (30.03%, *p*=0.03 in paired Wilcoxon test) and after 10 min (25.38%, *p*=0.03 in paired Wilcoxon test). Additionally, in the responder sub-group *β* power increase persists after the 10 minutes (1.77 fold relative to the baseline reading stage, *p*=0.03 in paired Student’s t-test, *p*=0.05 in paired Wilcoxon test). This suggests that improvement in speech quality may be mediated by activation of the *β*-frequency band in speech centers and the change spectral power of the *β* band may serve as a surrogate variable predictive of functional response in speech quality.

## DISCUSSION

In this study we assessed the response of adults with and without stuttering to auditory stimulation with binaural beats using a broad range of physiologic metrics.

### EEG

The hypothesis drove our efforts that participants with stuttering may exhibit lower *β*-power than controls [reference]. We were not able to confirm this in our cohort, potentially due to a low number of participants. However, we observed a hemispheric asymmetry in *β* activity in controls but not AWS, which is in agreement with observations in children who stutter^37^. Next, we were able to induce an increase of *β*-power during reading activity in AWS but not controls by means of auditory binaural beats stimulation. This did not create a significant left-right asymmetry in participants with stuttering. However, the increase in β-power in AWS was significant only in the left hemisphere on a more granular level. This is a desirable effect as the speech centers localize to the LTP junction (Wernicke) and LFT area (Broca). Further, we saw that upon auditory stimulation, the *β*-power increased in projections of both speech centers in AWS but not in controls.

### Results of stimulation in AWS: Speech

Next, we demonstrate that auditory stimulation is resulting in a speech quality improvement. The effectiveness of *β*-range BB stimulation to improve speech quality in AWS agrees with earlier studies suggesting a link between *β* deficit and stuttering ^6,25,41^. Moreover, our results suggest that there is a dose-effect relation of speech improvement on the increase of *β* power in speech centers. As a further step, we need to explore the effect of stimulation on a larger group of AWS with a longer follow-up.

### Results of stimulation in AWS: stress and well being

Stuttering is often accompanied by stress and neurosis^35^, which is another common target of stuttering therapy [[OASES](https://doi.org/10.1016/j.jfludis.2006.02.002)]. In this study, we investigated levels of stress by monitoring participants’ heart rate and heart rate variability (HRV). We observed a significant drop in stress index after exposure to the stimuli in participants with stuttering and controls, which may suggest a reduction of stress levels. This is in agreement with earlier work showing that brain waves can be entrained with BB^38,39^, which may cascade into other physiological functions such as HRV^36,^40.

### Further directions

As a further step, we plan to reproduce results with sham controls for stimuli (no-stimulation condition), and study the effect of both amplitude and the frequency of binaural shift (currently set to 21 Hz) on fluency improvements. Other tone frequencies should be further investigated because the optimal resonant frequency can be individual for different patients. The hypothesis of calibration of the target frequency may depend on individual brain electric activity and may be reflected in speech phonetic patterns. Event-related potentials (ERP) should be further analysed at the time when the stimulation occurs to evaluate the brain response to auditory sudden change, transitioning from environmental sound intensity to stimulus intensity, but the methodology should be time-fixed. Cross-over design and including the placebo trials of the experiment should be conducted to clarify effects of the stimulus that do not depend on groups of participants. Long-term effects of exposure, including possible habituation or long-term exposition, as well as impact on participants’ general well-being should be further investigated. The duration and effect of carrier music parameters as well as variations of the binaural shift frequency on the post-effect may be investigated in more detail in future research.

A longer stimulation may be also considered due to the non-invasiveness and absent side-effect of the proposed approach: Patient does not experience negative or irritating effects or fatigue after 5-minutes of BB stimulation. Thus, our novel auditory entrainment approach can be used as the multi-day course with several sessions.

### Potential applications

In-depth understanding of auditory BB stimulation effects could be useful for other kinds of end-user applications. For example, simple software applications for smartphones with built-in microphone and headphones with high-quality frequency characteristics could be used for usual daily exercises. This gives us a reason to believe that the fundamental results of current work can be used to create an original digital therapeutic technique for the treatment of speech disfluencies.

## Data Availability

All data produced in the present work are contained in the manuscript

https://drive.google.com/drive/folders/1h3Jg7hcAV5WNuRwLAHfTrzqOmTMIQLfS?usp=sharing

## ACKNOWLEDGMENTS

We are grateful to Dr. Elena Kruglyakova for productive scientific discussions and consultations on the neurophysiological aspects of speech production and EEG bio-signals processing.

## SUPPLEMENTARY MATERIALS

The Supplementary Materials including audio and video records demonstrating an effect of auditory stimulation can be found online at the following link: https://drive.google.com/drive/folders/1h3Jg7hcAV5WNuRwLAHfTrzqOmTMIQLfS?usp=sharing

---

**Supplementary Table 1.** EEG β-power values during the experiment, separately averaged for the electrodes of the left and right hemispheres.

| Condition | Cohort | Left ($\mu V^2$) | Right ($\mu V^2$) | p-value |
| --- | --- | --- | --- | --- |
| *Baseline reading* | AWS | 43.5 ± 21.9 | 54.8 ± 21.9 | 0.74 |
| *After stimulation* | AWS | 71.1 ± 42.1 | 53.8 ± 42.8 | 0.19 |
| *Post effect* | AWS | 67.4 ± 66.8 | 48.2 ± 61.0 | 0.38 |
| *Baseline reading* | controls | 39.9 ± 29.3 | 31.1 ± 26.1 | 0.03* |
| *After stimulation* | controls | 46.5 ± 36.6 | 53.5 ± 56.9 | 1.00 |
| *Post effect* | controls | 50.5 ± 64.1 | 28.6 ± 16.7 | 0.43 |

**Supplementary Figure 1.**
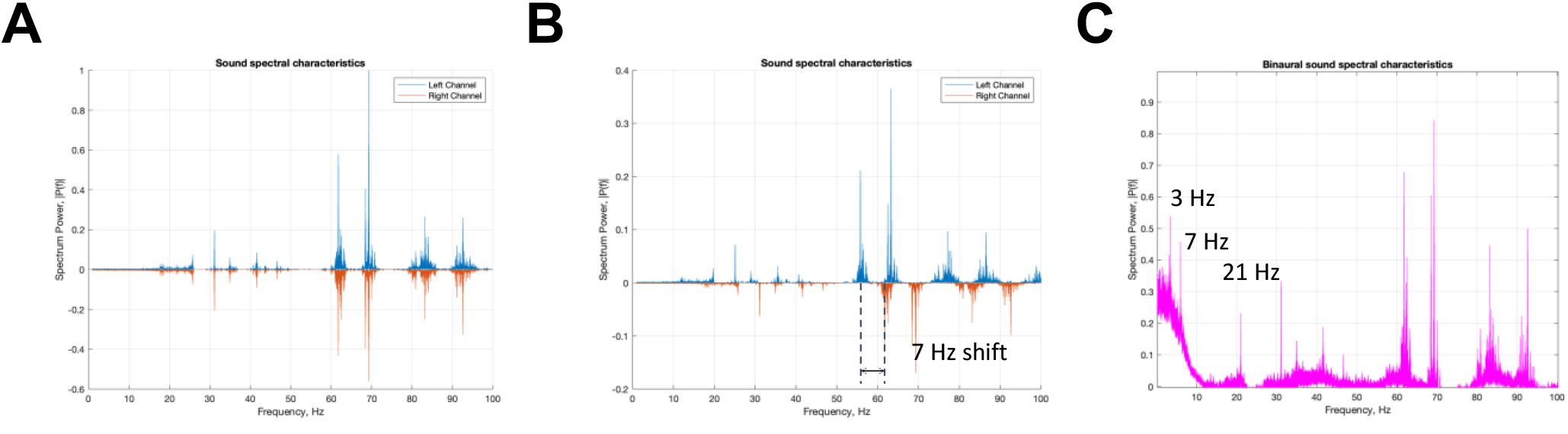
Spectral characteristics of auditory stimulus. **A**. Spectrum of an input signal. **B**. Spectrum of resulting signal with single target frequency (7 Hz) shift. **C**. Spectrum of output auditory stimulus as superposition of both channels with spectral components 3, 7 and 21 Hz.

**Supplementary Figure 2.**
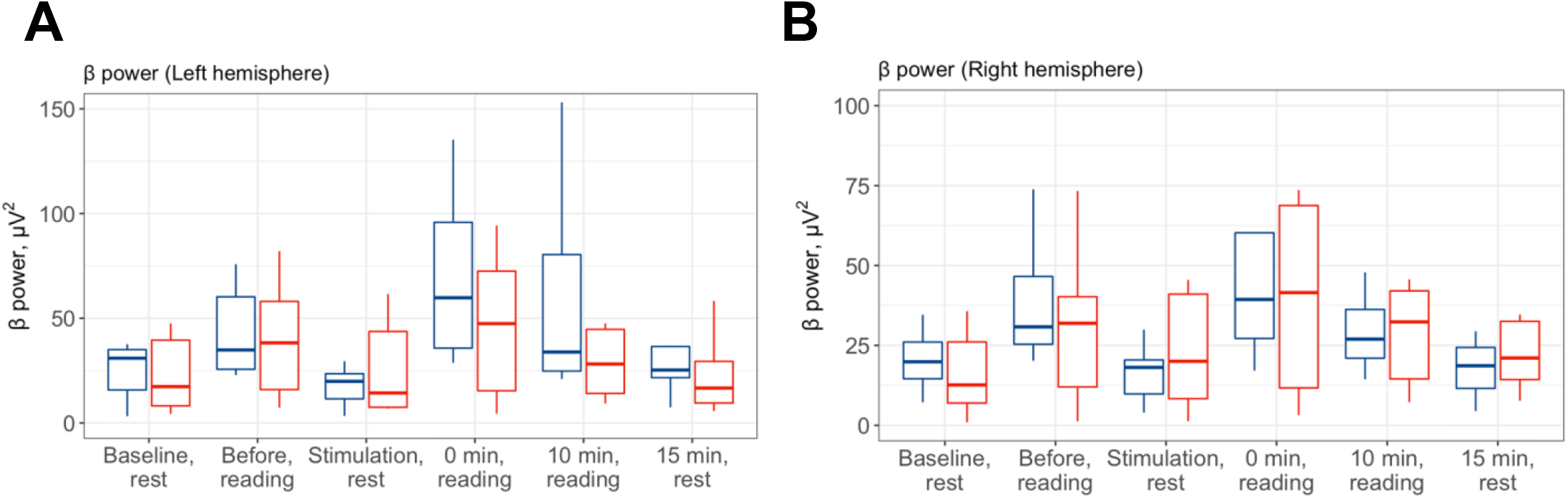
Changes in *β* power during the experiment. **A**. *β* power spectrum for left hemisphere in both cohorts during all stages of experiment. **B**. *β* power spectrum for right hemisphere in both cohorts during all stages of experiment.

**Supplementary Figure 3.**
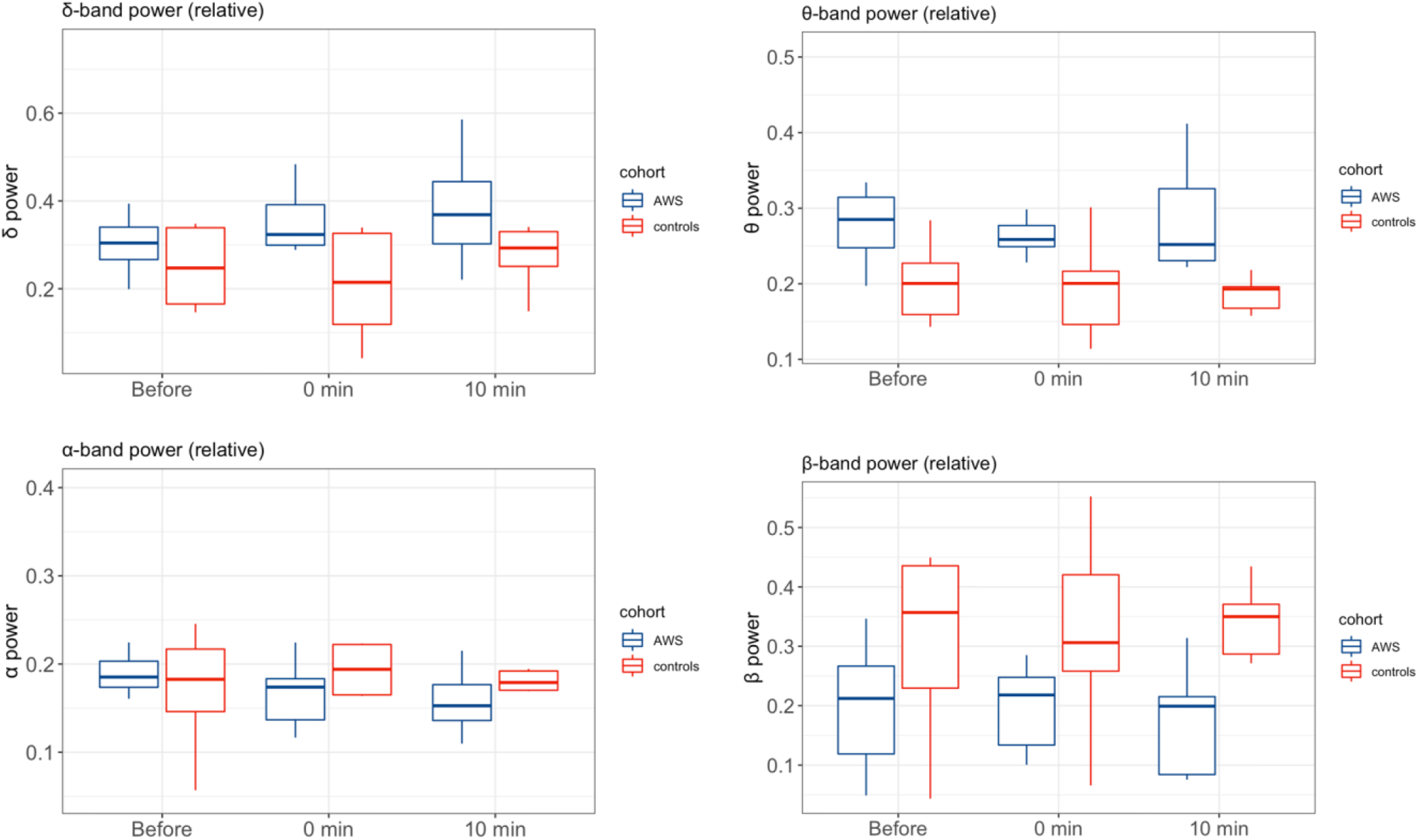
Changes in relative power averaged across all electrodes for different frequency bands (α, β, δ and θ) for reading tasks before, after and 10 minutes after binaural beats stimulation.

**Supplementary Figure 4.**
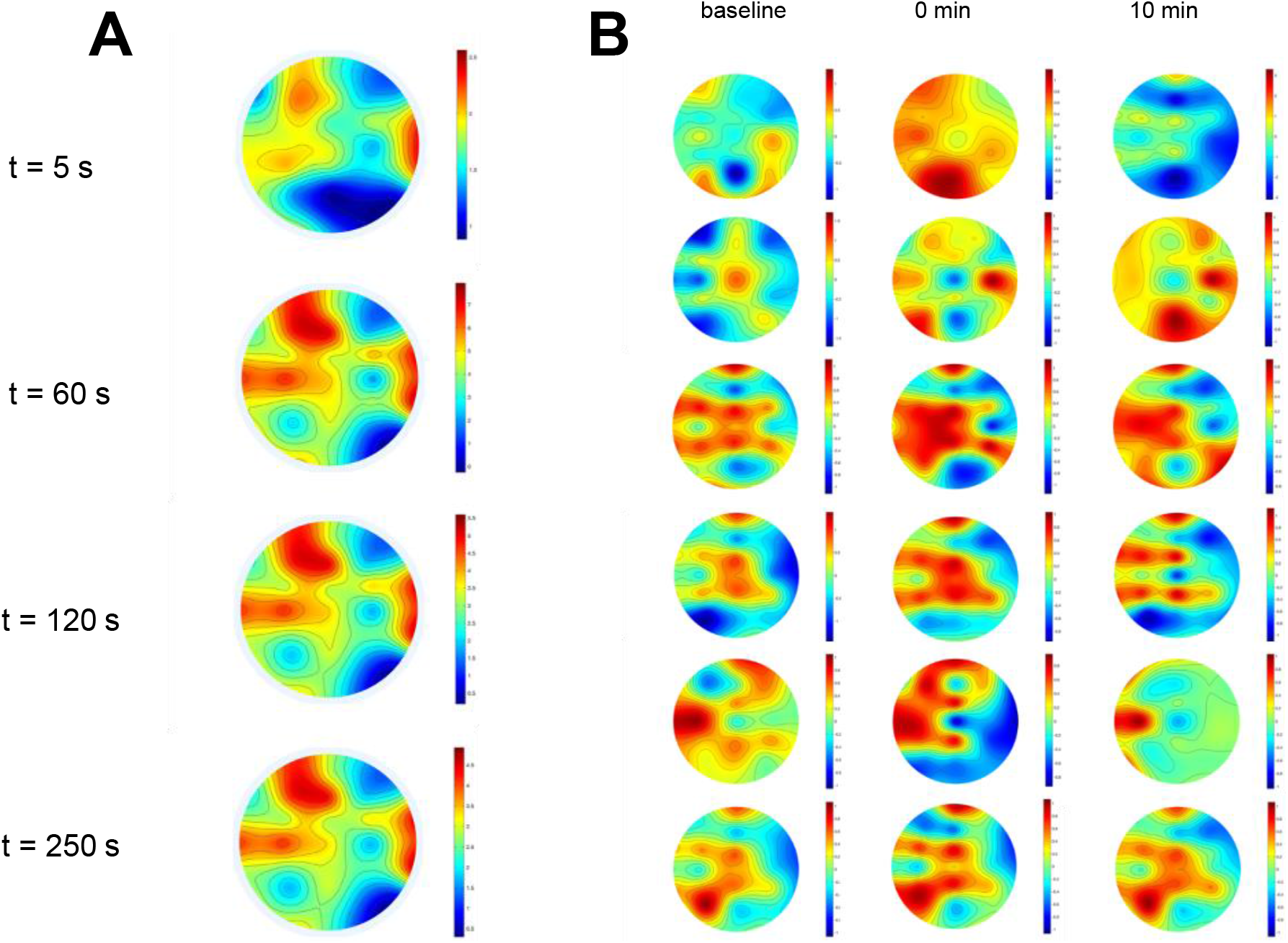
Time-resolved topography of *β-*power during binaural beats stimulation. **A**. From the top to bottom figures shown exposures of brain activity after 5, 60, 120 and 250 sec of stimulation beginning, respectively. **B**. Topography of *β*-power in reading experiments for each AWS participant (baseline, 0 min and 10 minutes after).

**Supplementary Figure 5.**
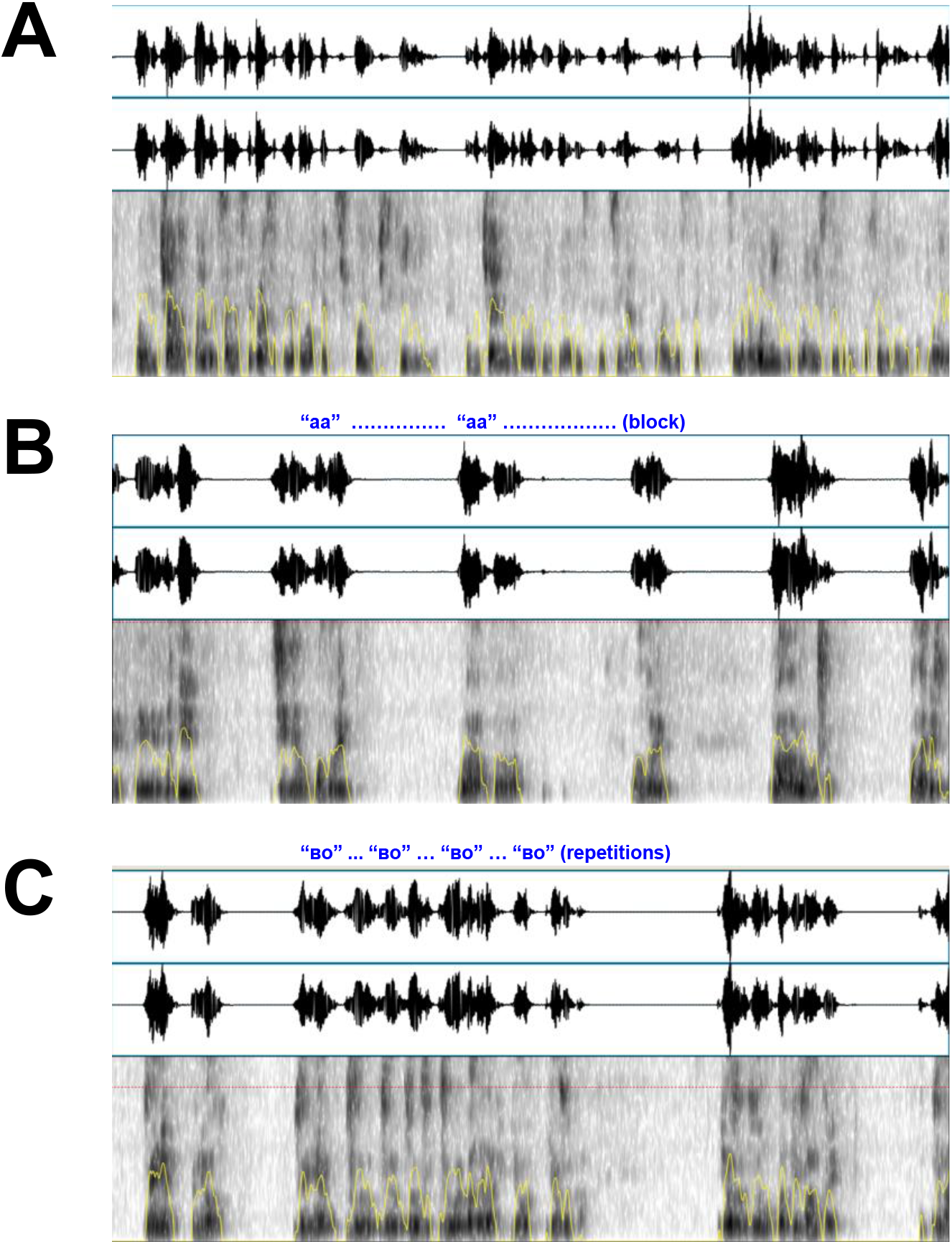
Snippets of the raw speech signals. Raw signal and spectral characteristics of fluent speech (**A**), speech with blocking episode (**B**) and repetitions episode (**C**).

**Supplementary Figure 6.**
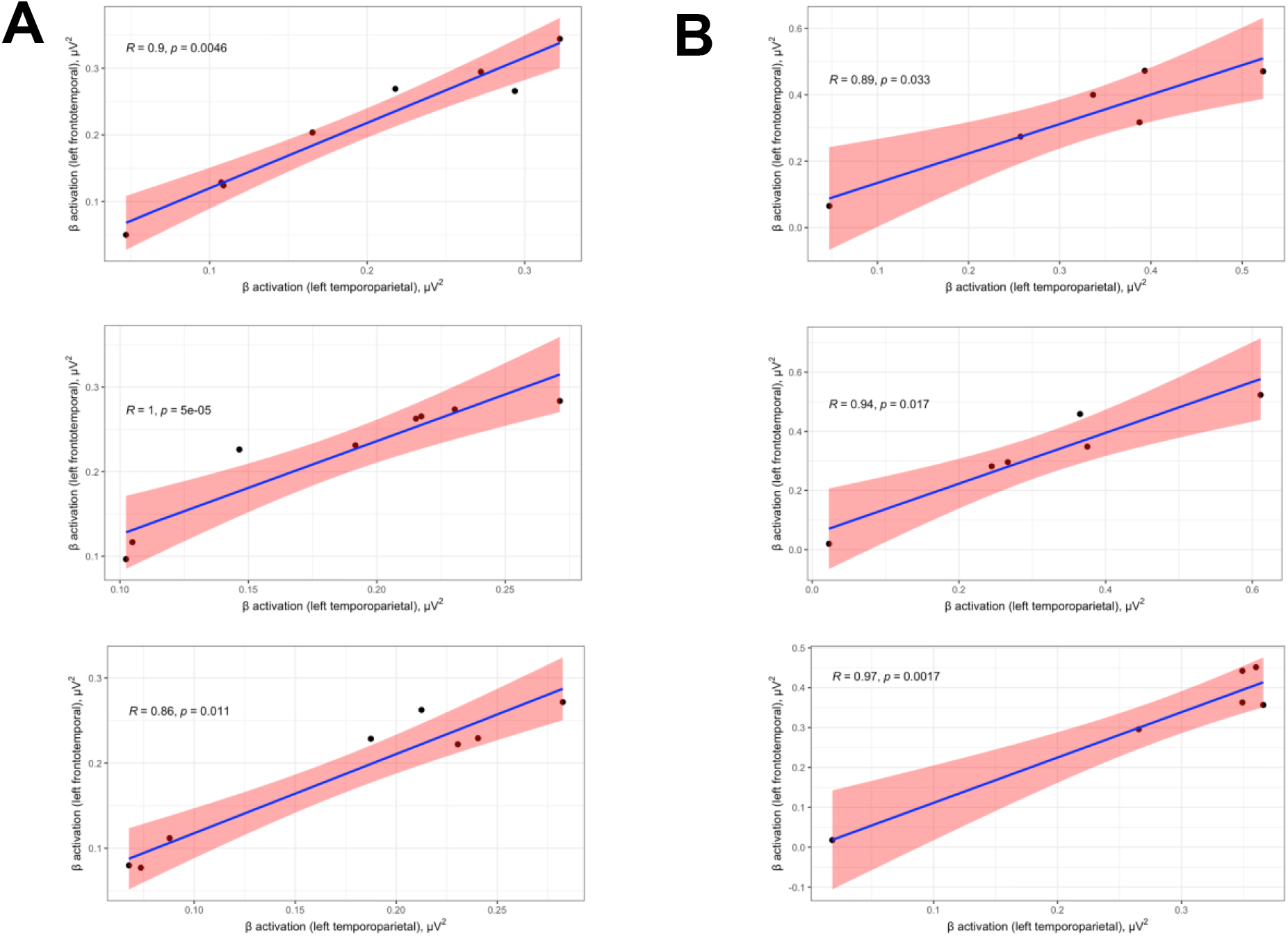
Cross-sectional correlation between normalized activity the left temporoparietal (LTP) projection and left frontotemporal (LFT) area in AWS (A) and controls (B) before (top), immediately after the stimulation (middle), and 10 minutes later (bottom).

**Supplementary Figure 7.**
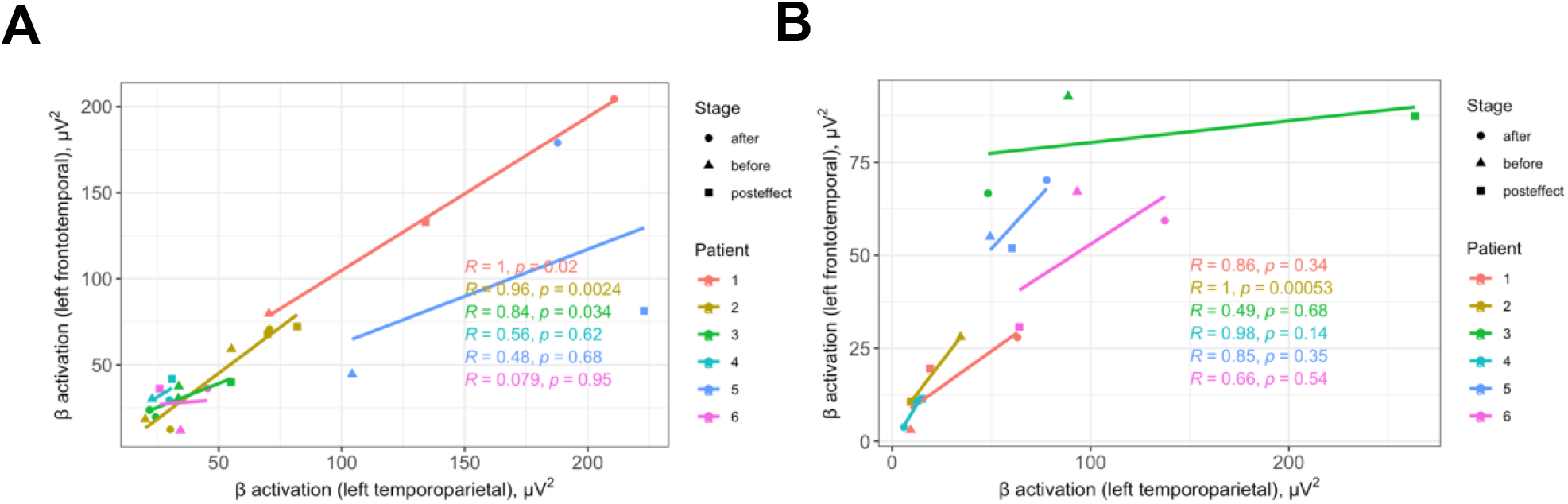
Cross-sectional correlation between activity the left temporoparietal (LTP) junction and left frontotemporal (LFT) area in AWS (A) and controls (B) for each AWS participant.

## Supplementary Figures

**Suppl. Table 1.**
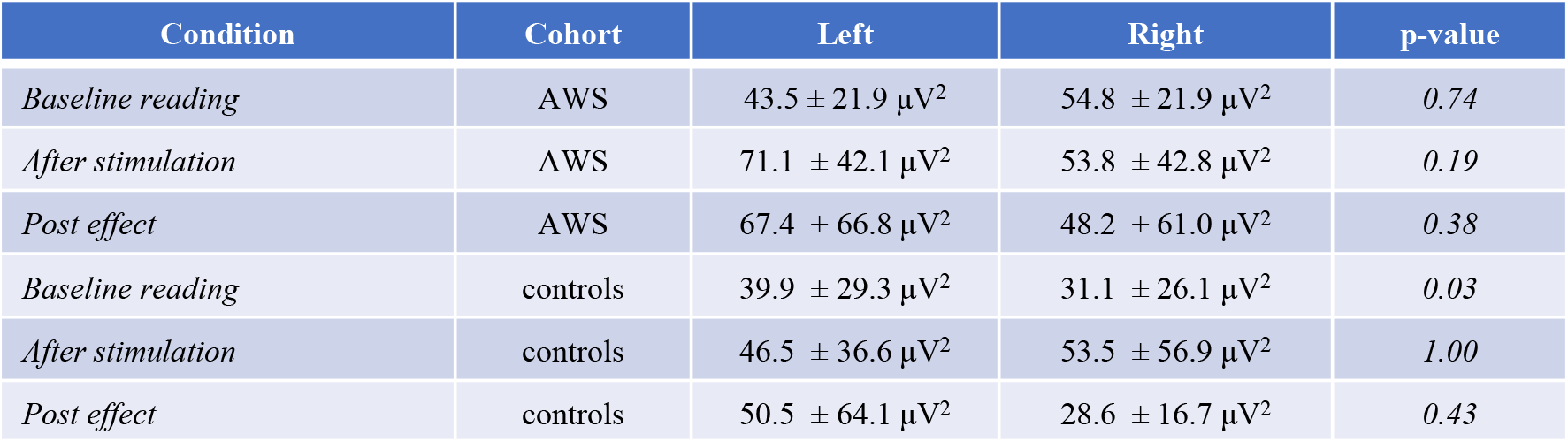
EEG β-waves power distribution between left and right hemispheres during the experiment.

